# Increasing severity, importance of viral transmission, and environmental exposures for pediatric asthma in a large urban health system, 2018-2023

**DOI:** 10.1101/2025.02.01.25321524

**Authors:** Sarah McCuskee, Rishi Kowalski, Cindy Mei, Alexis Zebrowski, Aman Patel, Lauren Zajac, Hashem Zikry, Ali Ayyash, Nicholas DeFelice

## Abstract

**Objective:** Pediatric asthma is multifactorial: social, environmental, and infectious exposures trigger exacerbations. Policies adopted to mitigate COVID-19 transmission in schools including school closures, mask mandates, and full reopening, provide a natural experiment to investigate the importance of socially-driven exposures, including viral transmission, in pediatric asthma.

**Methods:** This observational study used retrospective review of all electronic health records from the largest urban health system in New York City over five school years from 2018-2023. We investigated the epidemiology of acute, unscheduled care for pediatric asthma in children ages 5-17 years, including emergency department (ED) visits, hospital admissions, intensive care unit (ICU) stays, and viral testing. Comparison groups were drawn from citywide surveillance data and all-cause visits. We investigated population-level exposures: COVID-19 mitigation policies and wildfire smoke events.

**Results:** In the post-pandemic period, ED volume for pediatric asthma dropped (p<0.0001); however, children who presented were significantly more likely to require hospital admission and ICU care (p<0.001). Viral testing was employed more frequently, and more frequently positive, in the post-pandemic period (p<0.0001), with rhinovirus driving a greater proportion of pediatric asthma than all-cause adult or pediatric visits (p=0.02). After mask mandates were dropped in early 2022, pediatric asthma peaked during the immediate return-to-school fall period, slightly preceding the winter 2022 peak in viral illnesses. Wildfire smoke events were not significantly associated with pediatric asthma visits.

**Conclusions:** Socially-driven factors including viral transmission and school policies were important in driving pediatric asthma during and after the COVID-19 pandemic. Increased acuity despite lower volumes may help guide health systems as they strive to increase readiness to care for pediatric populations.

## Introduction

### Background

The pathogenesis of pediatric asthma is multifactorial and complex, but it is increasingly recognized as a socially-driven health condition.^1–3^. Exposure to respiratory viruses plays a substantial role in pathogenesis and exacerbation of symptoms, driving healthcare utilization and morbidity, but virus transmission is itself socially driven.^4^ Other etiologies, such as poor air quality including suspended particulate matter and aeroallergens, and triggers in the home environment including mold and other proinflammatory substances, are also socially driven^3^. These factors together are likely responsible for a significant proportion of the known geographic, income, and racial/ethnic variability in pediatric asthma prevalence and morbidity^5,6^.

The COVID-19 pandemic markedly changed several of these social drivers of pediatric asthma^4^. New York City (NYC) had a well-defined citywide “New York on PAUSE” policy including school closures, reopening with mandatory masking, and “demasking” (full opening without masking requirement). Previous literature has investigated pediatric asthma during the pandemic in parts of NYC^4^, but the timing and impact of school closures, the “New York on PAUSE” policy, school reopenings, and demasking have not been investigated. During the COVID-19 pandemic, testing for respiratory viruses also became commonplace in emergency departments.^2^ This provides a natural experiment which allows estimation of the impact of socially-driven respiratory virus transmission on pediatric asthma morbidity in a large urban population. Concomitantly, the NYC area experienced several unusually severe wildfire smoke events during the pandemic recovery period. Wildfire smoke has been associated with ill health outcomes, particularly from respiratory causes. Despite ecological data supporting this link, studies examining the impact of wildfire smoke on pediatric asthma are heterogeneous and lacked significant associations in a recent meta-analysis^7^. The pathophysiology and health system burden of wildfire smoke on pediatric asthma are incompletely understood.

### Importance

Understanding pediatric asthma epidemiology and healthcare utilization is critical to planning public health interventions. Undersampling of patients from groups exposed to multiple social drivers of disease, particularly in pediatric asthma, which is a racial disparity condition, may pose challenges for studies attempting to inform public health interventions and assess the overlapping impacts of multiple exposures. Therefore, we used observational data from the electronic health record (EHR) to mitigate selection biases and provide actionable estimates.^8^

### Goals of this investigation

We investigated the epidemiology, viral milieu, and environmental drivers of acute, unscheduled care for pediatric asthma across the largest health system in NYC in relation to the timing of policies intended to mitigate COVID-19 viral transmission over five school years, 2018-2023.

## Methods

### Study Design and Setting

This was a retrospective observational study of emergency department visits and inpatient hospital admissions for asthma in children ages 5-17 years, using routinely collected electronic health record data from 5 hospitals across Manhattan, Queens, and Brooklyn comprising the largest urban health system in New York City over the period 2018-2023.

The study was approved by the Institutional Review Board of the Icahn School of Medicine at Mount Sinai, STUDY-20-01285.

### Selection of Participants

To form the study population, we included all emergency department (ED), hospital, and intensive care unit (ICU) encounters for pediatric asthma during period 9/1/18 – 6/27/23. This period includes all full school years between 2018 and 2023. Asthma was defined using the primary encounter diagnosis code by International Classification of Diseases, 10^th^ edition (ICD-10) as codes ‘J45.20’, ‘J45.21’, ‘J45.22’, ‘J45.30’, ‘J45.31’, ‘J45.32’, ‘J45.40’, ‘J45.41’, ‘J45.42’, ‘J45.50’, ‘J45.51’, ‘J45.52’, ‘J45.901’, ‘J45.902’, ‘J45.909’, ‘J45.990’, ‘J45.991’, ‘J45.998’. The study population included children ages 5-17 at the time of each encounter. Analyses were conducted both by patient and by encounter, as some patients had repeat encounters. Hospital admissions and ICU stays occurring after transfer to the pediatric hospital from a different ED were linked using transfer data to the initial ED visit.

### Interventions

This was an observational study; however, we leveraged the natural experiment of changing policies directed at children in NYC during the study period. NYC public schools were closed on March 15, 2020. Schools fully reopened for in person instruction with a masking mandate in place on September 13, 2021. The masking mandate in schools was dropped effective March 8, 2022.

### Measurements

For each encounter, demographic (sex assigned at birth, insurance, patient guardian-declared race/ethnicity, zip code of residence), utilization variables (length of stay, ED/hospital/ICU admission, viral test administration), and clinical variables (viral test results) were collected. Two comparator populations were also extracted with the same variables as the study population: all ED visits in pediatric (ages 5-17 years) and all-age populations over the same period.

A third comparison dataset was created to assess representativeness of the detailed health system sample as well as infectious exposures. We extracted NYC Syndromic Surveillance data for the same periods, which indicates the total number of ED visits for either asthma or influenza-like illness (ILI+) in children ages 5-17. Data are provided by the NYC Department of Health and capture 100% of ED visits in the city.^9^ Visits are classified using text processing of chief complaints and ICD-10 discharge diagnosis codes. Asthma visits include ED visits among NYC resident children with ICD-10 principal diagnosis code of J45 or R06.2, or mention of “asthma, wheezing, or reactive airway”. ILI+ visits include mention of “flu, fever, and cough or sore throat”.

To assess environmental triggers for asthma, we extracted air quality data, pollen, and weather data. Fine particulate air pollution (PM2.5) were extracted from sensor data available on the US Environmental Protection Agency Air Quality System Data Mart.^10^ We took the mean reading from sensors closest in proximity to zip code tabulation areas represented in the patient dataset (list of sensors available in supplementary material). Using these data, we identified four wildfire smoke event periods using Air Quality Index (PM2.5>35.4 µg/m^3^/24h) from 2019 to 2023 in New York City and defined pre-and post-event 7-day periods. We extracted measured pollen counts (grains/m^3^) from Fordham University’s midtown Manhattan air sampler stratified by source (tree, grass and weed).^11^ Temperature and humidity were obtained by aggregating hourly values from NLDAS Meteorology Data averaged across 5 0.125 degree grid cells that encompass New York city.^12^

### Outcomes

The primary outcome was visit to the ED, inpatient hospital admission, or ICU admission in the selected study population. Secondary outcomes included viral testing utilization and results.

### Analysis

First, data were stratified by date, including by public school years from the 2018-19 school year to the 2022-23 school year, and by relevant school closure, mask mandate, and reopening dates. We compared groups using chi-squared, Fisher’s exact, or Kruskal-Wallis tests, as appropriate.

## Results

### Characteristics of study participants

This study included 4,311 visits for pediatric asthma across the five-year study period. Comparison groups included all all-age (N = 394,020) and pediatric (ages 5-17 years, N = 20,931) visits across the Mount Sinai Health System over the study period, and comprehensive syndromic surveillance data for health facilities in NYC.

### Main results

Visit volume in the study population was similar to NYC data for asthma syndrome visits, but differs from the NYC data for influenza-like illness (ILI+). As presented in Table 1, over the 5-year study period, ED visits for asthma declined, despite an increase after a precipitous drop during the 2019-20 school year, but did not rebound to their pre-pandemic baseline (p<0.0001). However, despite the overall decrease in ED visits for asthma in this population, hospital admissions and ICU stays increased during- and post-pandemic both in absolute terms, and as a proportion of ED visits (p<0.001). Viral testing was employed post-2020 with greater frequency (p<0.0001), and was more likely to be positive (p<0.0001). This trend was particularly prominent in children ages 5-17 with asthma, whereas all pediatric visits more closely tracked adult testing, as shown in Figure 1. There was no change in patient sex or race/ethnicity during the study period; in 97.3% of visits, the child was identified by parent/guardian as having a race or ethnicity other than White. We investigated sex differences in asthma presentations, as noted in Table 2. Overall, female children accounted for a greater proportion of ED visits for asthma, with no significant change in proportion over time. Hospital admissions were not significantly different between males and females over the study period. However, ICU admissions were higher in female children, particularly in the final year of the study period, 2022-23 (trend; p=0.05).

**Table 1:**
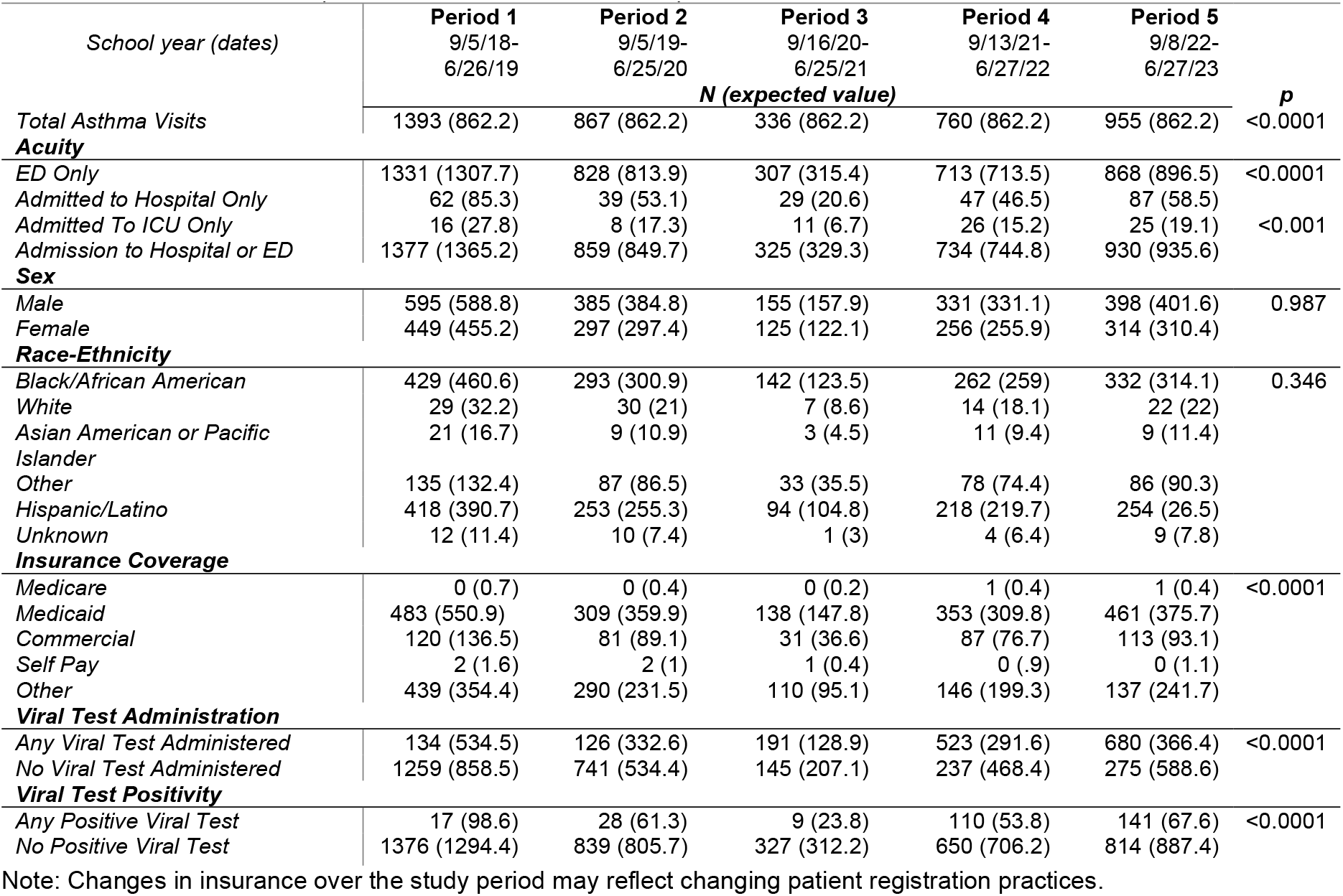
Pediatric Asthma Comparison Across School Years, ChiSq Tests.

| School year (dates) | Period 1<br>9/5/18-6/26/19 | Period 2<br>9/5/19-6/25/20 | Period 3<br>9/16/20-6/25/21 | Period 4<br>9/13/21-6/27/22 | Period 5<br>9/8/22-6/27/23 | *p* |
| --- | --- | --- | --- | --- | --- | --- |
|  | *N (expected value)* |  |  |  |  |  |
| **Total Asthma Visits** | 1393 (862.2) | 867 (862.2) | 336 (862.2) | 760 (862.2) | 955 (862.2) | <0.0001 |
| **Acuity** |  |  |  |  |  |  |
| ED Only | 1331 (1307.7) | 828 (813.9) | 307 (315.4) | 713 (713.5) | 868 (896.5) | <0.0001 |
| Admitted to Hospital Only | 62 (85.3) | 39 (53.1) | 29 (20.6) | 47 (46.5) | 87 (58.5) |  |
| Admitted To ICU Only | 16 (27.8) | 8 (17.3) | 11 (6.7) | 26 (15.2) | 25 (19.1) | <0.001 |
| Admission to Hospital or ED | 1377 (1365.2) | 859 (849.7) | 325 (329.3) | 734 (744.8) | 930 (935.6) |  |
| **Sex** |  |  |  |  |  |  |
| Male | 595 (588.8) | 385 (384.8) | 155 (157.9) | 331 (331.1) | 398 (401.6) | 0.987 |
| Female | 449 (455.2) | 297 (297.4) | 125 (122.1) | 256 (255.9) | 314 (310.4) |  |
| **Race-Ethnicity** |  |  |  |  |  |  |
| Black/African American | 429 (460.6) | 293 (300.9) | 142 (123.5) | 262 (259) | 332 (314.1) | 0.346 |
| White | 29 (32.2) | 30 (21) | 7 (8.6) | 14 (18.1) | 22 (22) |  |
| Asian American or Pacific Islander | 21 (16.7) | 9 (10.9) | 3 (4.5) | 11 (9.4) | 9 (11.4) |  |
| Other | 135 (132.4) | 87 (86.5) | 33 (35.5) | 78 (74.4) | 86 (90.3) |  |
| Hispanic/Latino | 418 (390.7) | 253 (255.3) | 94 (104.8) | 218 (219.7) | 254 (26.5) |  |
| Unknown | 12 (11.4) | 10 (7.4) | 1 (3) | 4 (6.4) | 9 (7.8) |  |
| **Insurance Coverage** |  |  |  |  |  |  |
| Medicare | 0 (0.7) | 0 (0.4) | 0 (0.2) | 1 (0.4) | 1 (0.4) | <0.0001 |
| Medicaid | 483 (550.9) | 309 (359.9) | 138 (147.8) | 353 (309.8) | 461 (375.7) |  |
| Commercial | 120 (136.5) | 81 (89.1) | 31 (36.6) | 87 (76.7) | 113 (93.1) |  |
| Self Pay | 2 (1.6) | 2 (1) | 1 (0.4) | 0 (.9) | 0 (1.1) |  |
| Other | 439 (354.4) | 290 (231.5) | 110 (95.1) | 146 (199.3) | 137 (241.7) |  |
| **Viral Test Administration** |  |  |  |  |  |  |
| Any Viral Test Administered | 134 (534.5) | 126 (332.6) | 191 (128.9) | 523 (291.6) | 680 (366.4) | <0.0001 |
| No Viral Test Administered | 1259 (858.5) | 741 (534.4) | 145 (207.1) | 237 (468.4) | 275 (588.6) |  |
| **Viral Test Positivity** |  |  |  |  |  |  |
| Any Positive Viral Test | 17 (98.6) | 28 (61.3) | 9 (23.8) | 110 (53.8) | 141 (67.6) | <0.0001 |
| No Positive Viral Test | 1376 (1294.4) | 839 (805.7) | 327 (312.2) | 650 (706.2) | 814 (887.4) |  |

Note: Changes in insurance over the study period may reflect changing patient registration practices.

**Table 2:** Comparison of Sex and Admission Status, MSSM Asthma Presentations Across School Years, Chi-squared tests.

| School year (dates) | Period 1 | Period 2 | Period 3 | Period 4 | Period 5 |  |
| --- | --- | --- | --- | --- | --- | --- |
|  | 9/5/18-6/26/19 | 9/5/19-6/25/20 | 9/16/20-6/25/21 | 9/13/21-6/27/22 | 9/8/22-6/27/23 |  |
| **ED Only Visits** | **N (expected value)** |  |  |  |  | **p** |
| Sex Male | 566 (586.3) | 356 (353.5) | 130 (131.1) | 303 (304.4) | 373 (370.6) | 0.999 |
| Sex Female | 765 (762.7) | 472 (474.6) | 177 (175.9) | 410 (408.6) | 495 (497.4) |  |
| **Admitted to Hospital** | **N (expected value)** |  |  |  |  | **p** |
| Sex Male | 21 (23) | 17 (15.5) | 9 (9) | 10 (10.5) | 32 (31) | 0.945 |
| Sex Female | 25 (23) | 14 (15.5) | 9 (9) | 11 (10.5) | 30 (31) |  |
| **ICU Admission** | **N (expected value)** |  |  |  |  | **p** |
| Sex Male | 8 (8.2) | 4 (4.1) | 10 (5.6) | 13 (13.3) | 9 (12.8) | 0.0544 |
| Sex Female | 8 (7.8) | 4 (3.9) | 1 (5.4) | 13 (12.7) | 16 (12.2) |  |

**Fig 1.**
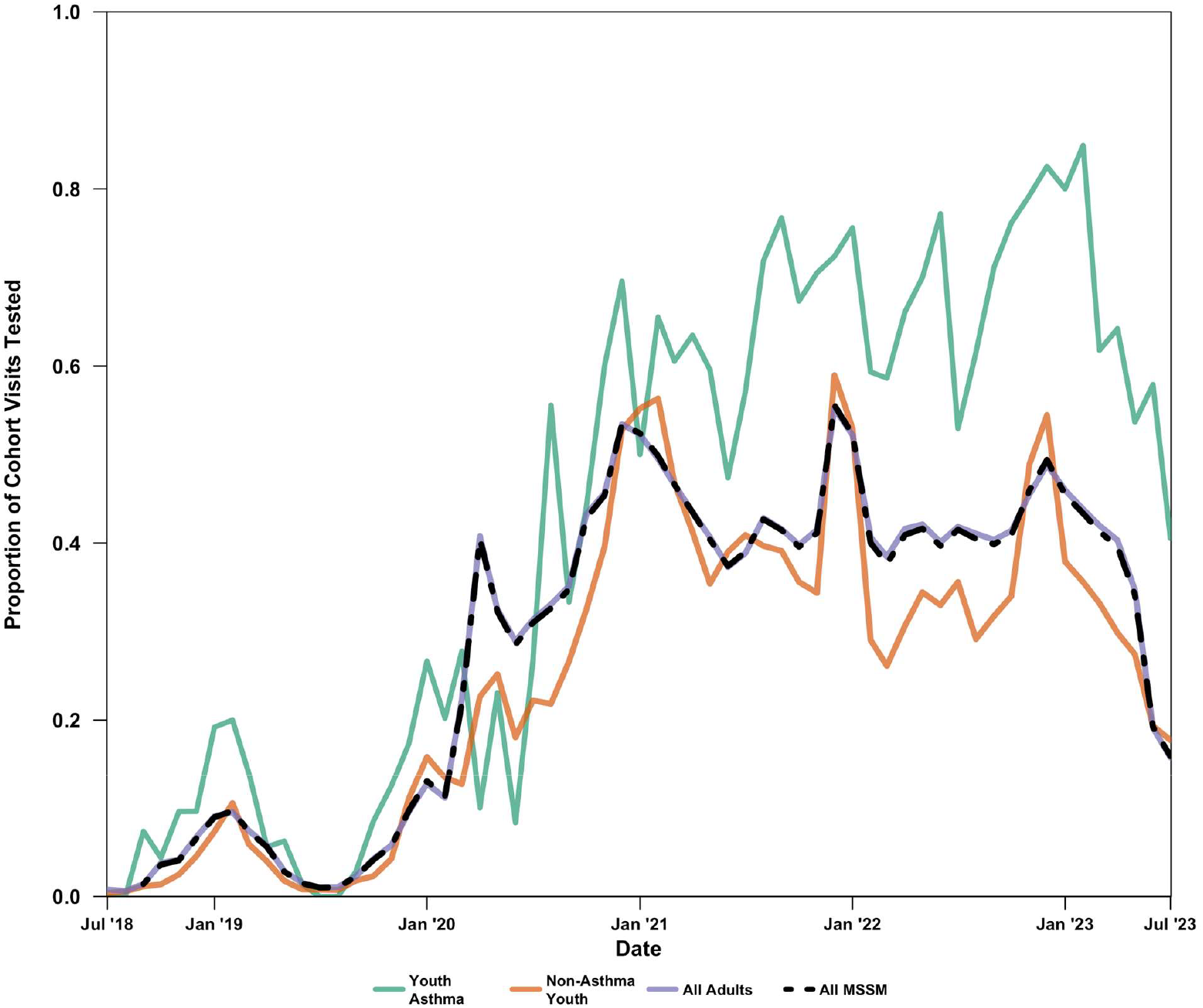
Monthly Proportion of Patients Administered Any Viral Test by Patient Cohort, July 2018 – July 2023.

Need for critical care for asthma increased in the post-pandemic period. Table 3 compares patients with asthma diagnoses admitted to ICU to all ICU admissions in patients 5-17 years old. Overall, there was an increase in the number of children admitted to ICU and in the proportion of those children admitted for asthma (p<0.0001). All children admitted to ICU were tested for at least one virus; the proportion of those testing positive increased over the study period, peaking in the 2021-2022 school year with 84.6% of patients admitted to ICU for asthma testing positive for a virus (p<0.0001). Among ICU patients, there were no differences in gender, race/ethnicity, or insurance over the study period. Overall, as shown in Table 4, ICU length of stay was shorter for patients with asthma across all five years of the study (mean 137.5 (standard deviation, SD 301.4) hours vs. mean 42.7 (SD 26.2) hours, p<0.0001). ICU patients who tested positive for a virus had longer lengths of stay, both for non-asthma (p=0.03) and a trend for asthma ICU patients (p=0.08). In school years 2021-2022 (period 4) and 2022-2023 (period 5), increased viral positivity coincided with overall longer lengths of stay in patients admitted to the hospital (supplementary figure 2).

**Table 3.**
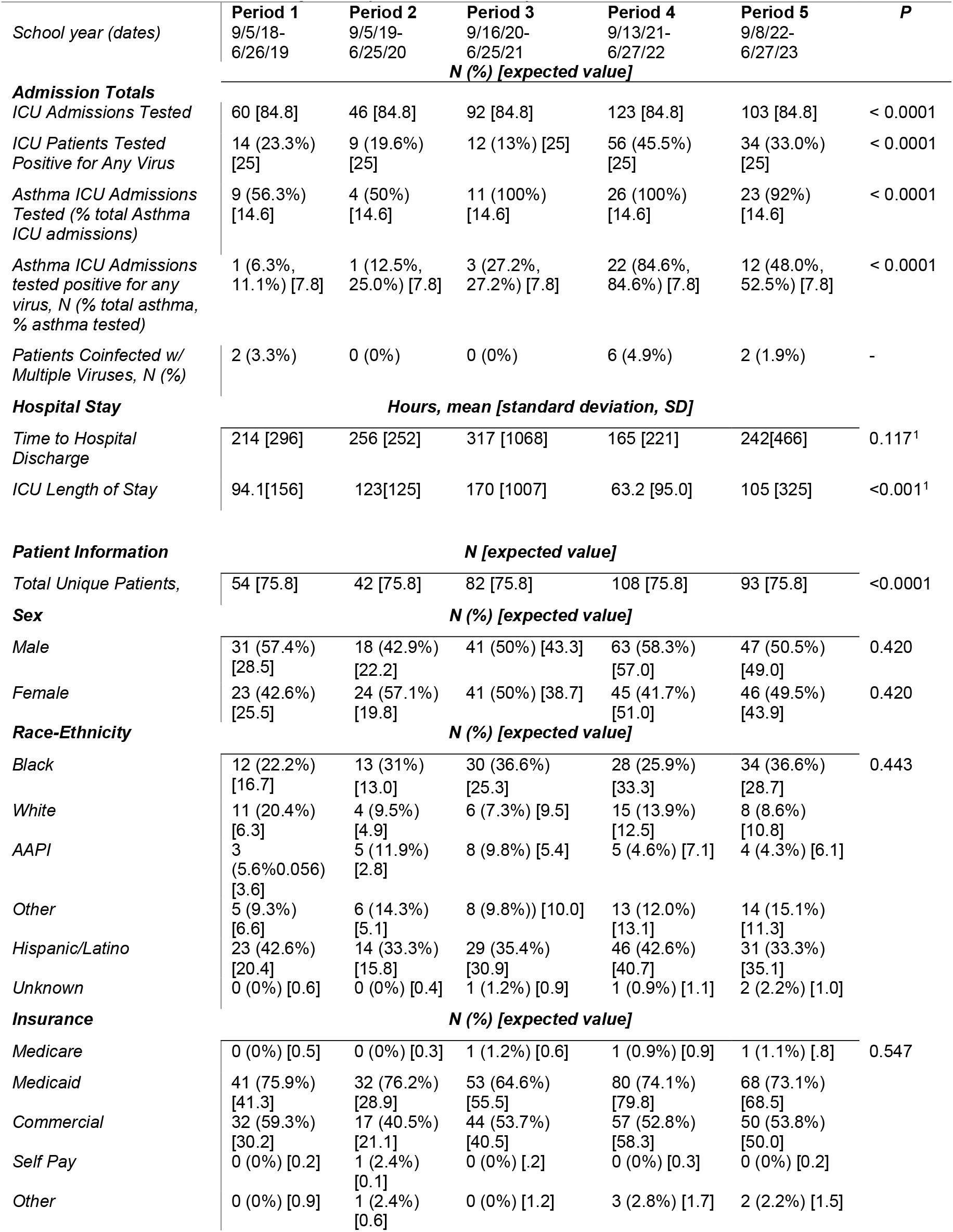
ICU Admissions in children ages 5-17 years across school years.

| School year (dates) | Period 1<br>9/5/18-6/26/19 | Period 2<br>9/5/19-6/25/20 | Period 3<br>9/16/20-6/25/21 | Period 4<br>9/13/21-6/27/22 | Period 5<br>9/8/22-6/27/23 | P |
| --- | --- | --- | --- | --- | --- | --- |
| **Admission Totals** |  |  |  |  |  |  |
| ICU Admissions Tested | 60 [84.8] | 46 [84.8] | 92 [84.8] | 123 [84.8] | 103 [84.8] | < 0.0001 |
| ICU Patients Tested Positive for Any Virus | 14 (23.3%) [25] | 9 (19.6%) [25] | 12 (13%) [25] | 56 (45.5%) [25] | 34 (33.0%) [25] | < 0.0001 |
| Asthma ICU Admissions Tested (% total Asthma ICU admissions) | 9 (56.3%) [14.6] | 4 (50%) [14.6] | 11 (100%) [14.6] | 26 (100%) [14.6] | 23 (92%) [14.6] | < 0.0001 |
| Asthma ICU Admissions tested positive for any virus, N (% total asthma, % asthma tested) | 1 (6.3%, 11.1%) [7.8] | 1 (12.5%, 25.0%) [7.8] | 3 (27.2%, 27.2%) [7.8] | 22 (84.6%, 84.6%) [7.8] | 12 (48.0%, 52.5%) [7.8] | < 0.0001 |
| Patients Coinfected w/ Multiple Viruses, N (%) | 2 (3.3%) | 0 (0%) | 0 (0%) | 6 (4.9%) | 2 (1.9%) | - |
| **Hospital Stay** |  |  |  |  |  |  |
| Time to Hospital Discharge | 214 [296] | 256 [252] | 317 [1068] | 165 [221] | 242[466] | 0.117<sup>1</sup> |
| ICU Length of Stay | 94.1[156] | 123[125] | 170 [1007] | 63.2 [95.0] | 105 [325] | <0.001<sup>1</sup> |
| **Patient Information** |  |  |  |  |  |  |
| Total Unique Patients, | 54 [75.8] | 42 [75.8] | 82 [75.8] | 108 [75.8] | 93 [75.8] | <0.0001 |
| **Sex** |  |  |  |  |  |  |
| Male | 31 (57.4%) [28.5] | 18 (42.9%) [22.2] | 41 (50%) [43.3] | 63 (58.3%) [57.0] | 47 (50.5%) [49.0] | 0.420 |
| Female | 23 (42.6%) [25.5] | 24 (57.1%) [19.8] | 41 (50%) [38.7] | 45 (41.7%) [51.0] | 46 (49.5%) [43.9] | 0.420 |
| **Race-Ethnicity** |  |  |  |  |  |  |
| Black | 12 (22.2%) [16.7] | 13 (31%) [13.0] | 30 (36.6%) [25.3] | 28 (25.9%) [33.3] | 34 (36.6%) [28.7] | 0.443 |
| White | 11 (20.4%) [6.3] | 4 (9.5%) [4.9] | 6 (7.3%) [9.5] | 15 (13.9%) [12.5] | 8 (8.6%) [10.8] |  |
| AAPI | 3 (5.6%) [3.6] | 5 (11.9%) [2.8] | 8 (9.8%) [5.4] | 5 (4.6%) [7.1] | 4 (4.3%) [6.1] |  |
| Other | 5 (9.3%) [6.6] | 6 (14.3%) [5.1] | 8 (9.8%) [10.0] | 13 (12.0%) [13.1] | 14 (15.1%) [11.3] |  |
| Hispanic/Latino | 23 (42.6%) [20.4] | 14 (33.3%) [15.8] | 29 (35.4%) [30.9] | 46 (42.6%) [40.7] | 31 (33.3%) [35.1] |  |
| Unknown | 0 (0%) [0.6] | 0 (0%) [0.4] | 1 (1.2%) [0.9] | 1 (0.9%) [1.1] | 2 (2.2%) [1.0] |  |
| **Insurance** |  |  |  |  |  |  |
| Medicare | 0 (0%) [0.5] | 0 (0%) [0.3] | 1 (1.2%) [0.6] | 1 (0.9%) [0.9] | 1 (1.1%) [0.8] | 0.547 |
| Medicaid | 41 (75.9%) [41.3] | 32 (76.2%) [28.9] | 53 (64.6%) [55.5] | 80 (74.1%) [79.8] | 68 (73.1%) [68.5] |  |
| Commercial | 32 (59.3%) [30.2] | 17 (40.5%) [21.1] | 44 (53.7%) [40.5] | 57 (52.8%) [58.3] | 50 (53.8%) [50.0] |  |
| Self Pay | 0 (0%) [0.2] | 1 (2.4%) [0.1] | 0 (0%) [0.2] | 0 (0%) [0.3] | 0 (0%) [0.2] |  |
| Other | 0 (0%) [0.9] | 1 (2.4%) [0.6] | 0 (0%) [1.2] | 3 (2.8%) [1.7] | 2 (2.2%) [1.5] |  |

**Table 4.** ICU Length of Stay (LOS) By Test Positivity and Asthma Diagnosis.

|  | LOS, Mean Hours<br>[SD] | P<sup>1</sup> |
| --- | --- | --- |
| **Non-Asthma ICU Admissions** |  |  |
| *All* | 138 [301] |  |
| *No Positive Viral Test* | 90.8 [221] | <0.05 |
| *Any Positive Viral Test* | 220 [1050] |  |
| **Asthma ICU Admissions** |  |  |
| *All* | 42.7 [26.2] |  |
| *No Positive Viral Test* | 30.0 [23.9] | 0.08 |
| *Any Positive Viral Test* | 43.2 [31.6] |  |

1 Kruskal-Wallis Test

Calendar year 2022 provides a natural experiment on the impact of masking in schools. NYC public schools required their students to wear a mask until March 8^th^ 2022, when the policy shifted to masks optional. In September 2022, schools reopened for the first school year without restrictions related to COVID-19. We plotted ED visits from NYC-wide syndromic surveillance data for asthma and ILI+ and Mount Sinai ED visits for (MSSM) asthma in the 5-17 years age group in Figure 2A. Seasonal variation is expected, as shown in Figure 2B, which demonstrates lower visits for both asthma and ILI+ during non-school year periods (dashed lines). The fall 2022 peak in asthma slightly preceded the winter 2022 peak in ILI+, occurring almost immediately during the return-to-school period (Figure 2).

**Figure 2.**
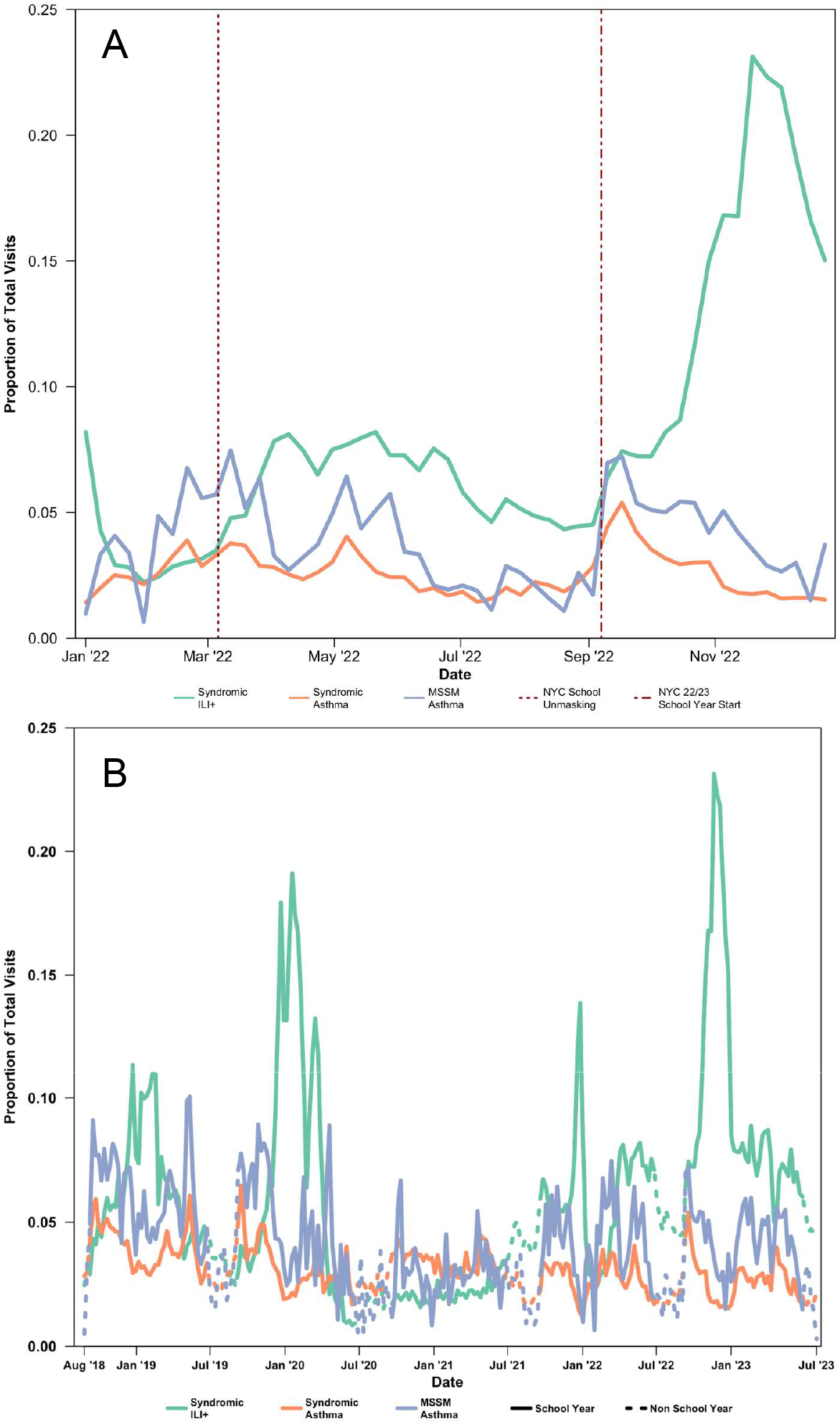
Weekly Proportions of MSSM Asthma and NYC Syndromic Admissions, Ages 5-17, A: Year 2022. B: 2018-2023.

Viral triggers for asthma differed from viral triggers for all-cause ED visits and hospitalizations in both the pediatric and all-age health system comparator populations. (Figure 3). COVID-19 and influenza A and B accounted for large proportions of both pediatric and all-age ED visits and hospitalizations in all comparator school year periods. However, uniquely in pediatric asthma visits, rhinovirus appeared to be an important contributor in every year after 2019, comprising 34-67% of viruses identified in pediatric asthma patients each year, while never accounting for more than 10% of all-cause adult and pediatric visits (p=0.02).

**Figure 3.**
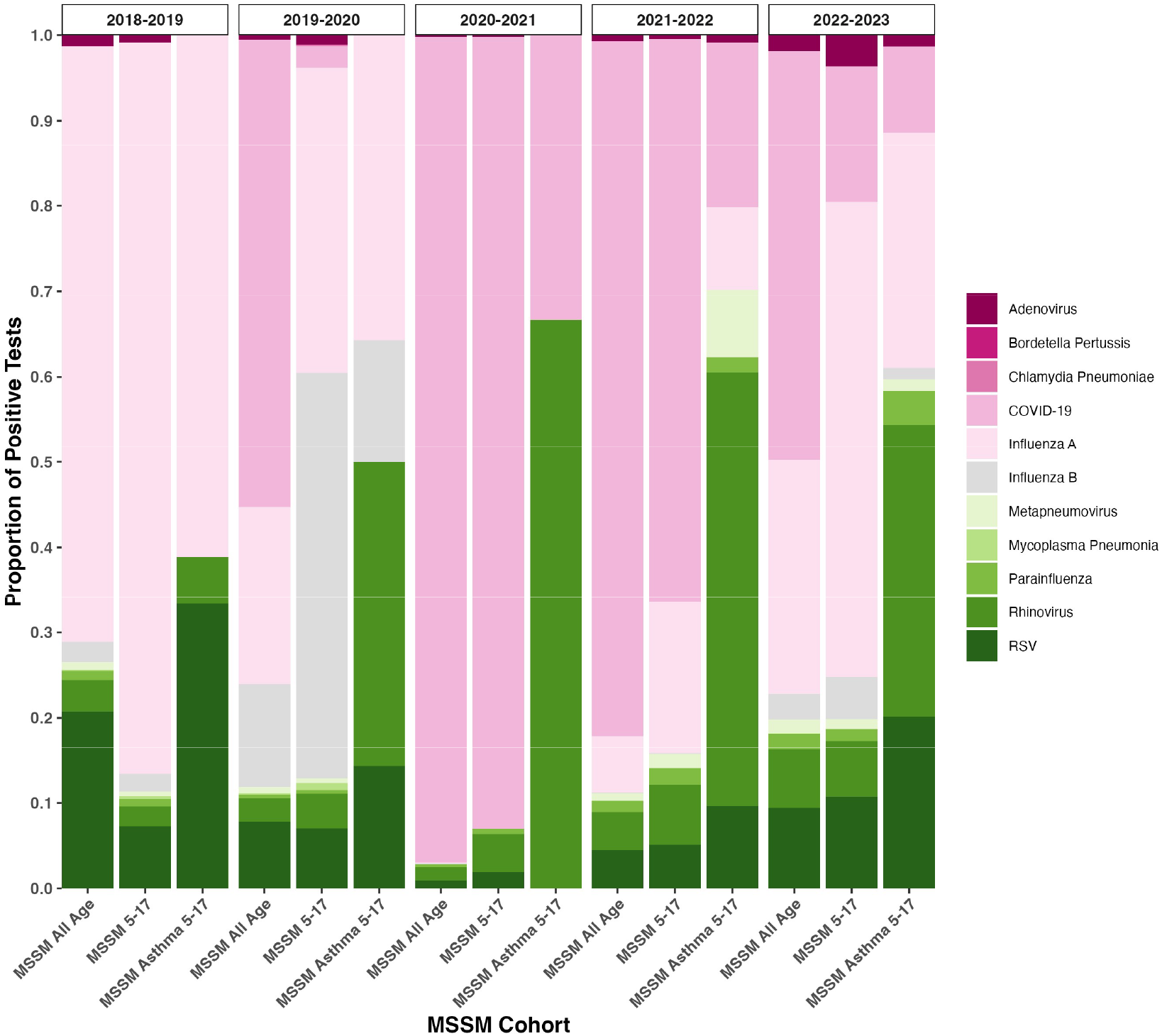
Comparison of Viral Testing Mixture between Pediatric Asthma Visits (ages 5-17 years), All Pediatric Visits, and All-Age Visits, Mount Sinai (MSSM), by School Year.

Finally, wildfire smoke poses an additional risk for children with asthma. In NYC during the study period, wildfire smoke events occurred near or after the end of the school year. Multiple exposures vary on a seasonal basis, including viral milieu, environmental triggers, and indoor allergens.^13^ In the pediatric asthma patients presenting to Mount Sinai, there was no statistically significant difference between the 7 days pre-event, the event period, and the 7 days post-event in ED visit rates (Table 5). There was a trend towards lower daily rates of ED visits for asthma (p=0.08) and a finding of lower visits for ILI+ (p<0.05) in the during- and post-event periods in NYC-wide syndromic surveillance data. Grass pollen was lower in the post-event period (p<0.05). Gender differences were observed in post-wildfire asthma presentations (70.7% male, vs. pre-event 42.6% male, p<0.01). No race/ethnicity differences were observed.

**Table 5:** Selected Exposure and Demographic Variables for Mount Sinai Asthma Admissions Pre, During and After Four Wildfire-related Acute Air Quality Events in NYC.

|  | Pre-Exposure | Exposure<br>Mean [SD] | Post-Exposure | P |
| --- | --- | --- | --- | --- |
| **Airborne exposures** |  |  |  |  |
| Daily PM2.5 Air Quality Index, $\mu\text{g}/\text{m}^3$ | 42.9 [12.9] | 144.7 [54.5] | 58.5 [21.1] | <0.0001 |
| Daily Tree Pollen Count | 27.1 [41.9] | 64.7 [78.6] | 8.07 [6.11] | 0.52 |
| Daily Grass Pollen Count | 3.65 [4.5] | 6.79 [5.7] | 1.91 [2.56] | <0.05 |
| Daily Weed Pollen Count | 2.31 [3.27] | 1.28 [2.07] | 1.27 [2.08] | 0.56 |
| **NYC Syndromic Surveillance** |  |  |  |  |
| Daily NYC All-Age Visits for Asthma | 158 [20.6] | 203 [71.1] | 162 [31.0] | 0.20 |
| Daily NYC All-Age Visits for ILI+ * | 208 [52.7] | 212 [33.2] | 204 [37.6] | 0.80 |
| **MSSM Non-Asthma Visits, ages 5-17 years** |  |  |  |  |
| Daily ED Visits | 14.0 [6.12] | 11.6 [5.83] | 12.1 [6.19] | 0.47 |
| Daily ED Visits with Positive Viral Test | 1.29 [1.18] | 0.71 [1.11] | 1.46 [1.20] | 0.26 |
| **MSSM Asthma Visits, ages 5-17 years** |  |  |  |  |
| Daily Visits (ED, Hospital, ICU) | 1.82 [1.93] | 2.29 [2.14] | 1.50 [1.37] | 0.64 |
| ED Visits | 1.68 [1.74] | 2.29 [2.14] | 1.39 [1.23] | 0.59 |
| Daily Visits (ED, Hospital, ICU), Positive Viral Test | 0.107 [0.32] | 0.14 [0.38] | 0.071 [0.26] | 0.81 |
| **Sex** |  |  |  |  |
|  | N (%) |  |  |  |
| Male | 20 (42.6) | 5 (31.2) | 29 (70.7) | <0.01 |
| Female | 27 (57.4) | 11 (68.8) | 12 (29.3) |  |
| **Race-Ethnicity** |  |  |  |  |
|  | N (%) |  |  |  |
| NH Black | 23 (48.9) | 9 (56.2) | 20 (48.8) | 0.69 |
| NH White | 1 (2.1) | 1 (6.2) | 0 (0) |  |
| NH AAPI | 0 (0) | 1 (6.2) | 2 (4.9) |  |
| NH Other | 4 (8.5) | 1 (6.2) | 4 (9.8) |  |
| Hispanic/Latino | 19 (40.4) | 4 (25.0) | 14 (34.1) |  |
| Unknown | 0 (0) | 0 (0) | 1 (2.4) |  |

\* ILI+: Viral syndromes, as described in text.

### Limitations

This observational study has several limitations. Although using routine health system data allowed us to minimize inclusion biases, differences in medical coding of asthma visits when driven by a viral infection may contribute to misclassification bias in patients with asthma if a visit for asthma exacerbation had a principal diagnosis code related to the viral etiology.

However, NYC-wide syndromic surveillance ILI+ data, which uses free-text search in addition to ICD codes, showed a very similar pattern to our cohort, suggesting that the risk of this misclassification may be minimal. Secondly, we did not observe racial/ethnicity changes over time during our study period in our study population. This may reflect the largely minoritized study population. This proportion varies substantially in the literature across geographic areas^14^, but appears representative based on other literature from New York City (cite). Differences in gender should be further explored^7^, as should geographic and systematic disparities which have been repeatedly demonstrated in existing literature.

## Discussion

This study underscores the importance of respiratory viral transmission as one of multiple dynamic, seasonal risks in pediatric asthma using the natural experiment of the COVID-19 pandemic and the policies intended to mitigate it. Despite lower total ED volume for asthma in the pediatric population, those children who did present were significantly more likely to require hospital admission and ICU care. Multiple factors may contribute to this, including changes in care-seeking behavior during and post-pandemic, changes in telemedicine or home management strategies undertaken by healthcare professionals, such as nebulizer or steroid prescriptions enabling longer self-management of exacerbations, and differential exposures^15^. COVID-19 itself did not drive a majority of pediatric asthma presentations in any school year across the study period. However, positivity for any virus in ICU patients was associated with longer length of stay in both asthma and non-asthma presentations.

As expected, COVID-19 and influenza A and B drove a majority of all-age and pediatric ED visits. However, viral triggers in the pediatric asthma population were distinct, particularly the importance of rhinovirus. This has been previously documented in other literature^16,17^; however its persistence as an important driver during both COVID-19 surges and the winter 2022 COVID/influenza/respiratory syncytial virus (RSV) surge is notable. The relative lack of rhinovirus detection in 2018-2019 may reflect the more limited testing strategy employed prior to the COVID-19 pandemic (Figure 1 and Table 1).

Policies such as school closure/reopening and masking/demasking, intended to mitigate COVID-19 spread, appeared to have significant associations with pediatric asthma presentations, possibly driven by respiratory virus transmission. After school mask mandates were dropped in spring 2022, there were few pediatric asthma presentations during the summer 2022. However, the return to school in fall 2022 was accompanied by surges in asthma, followed by ILI+. The timing of these two peaks may reflect the increased vulnerability of the pediatric asthma population to exposures including viral transmission in schools, or unmeasured competing risks, although contributors including particulate air pollution, PM2.5, temperature, humidity, and pollen count did not appear substantially different during this period (Supplementary Figure 1).

Clinical practice in application of viral testing, especially multiplex viral testing, became more liberal during the study period^18^, which is likely to have influenced the distribution of viruses detected from 2018 to 2023. However, pediatric asthma patients had distinct viral milieu within each school year when compared to the general pediatric and all-age health system populations. Pediatric asthma has many potential triggers which vary seasonally and in response to policies and unexpected events including pollution sources such as wildfires, traffic emissions, residential combustion, and industrial processes. In contrast to some prior studies^19^, but in keeping with others^20^, our research did not find significant differences in pediatric asthma or ILI+ presentations in the time periods pre-, during-, and post-wildfire smoke events. This may reflect the sensitivity of pediatric asthma to different choices of reference periods given the multiple competing exposures present, or changes in behavior during highly publicized wildfire smoke events. Other literature has demonstrated that wildfires are associated with increases in viral transmission in all-age populations^21^, a mixture of exposures which should be investigated in pediatric asthma.

Finally, our finding that female children were more likely to have presentations for asthma differs from much existing literature.^16,17^ One large study found that a history of pneumonia was more common in girls with asthma than boys with asthma^17^; given the importance of infectious (viral) triggers in our cohort, further research could investigate sex differences in relation to infectious asthma exacerbation triggers.

In summary, this study underscores the importance of policy decisions in influencing the complex mixture of social and environmental exposures that drive pediatric asthma in a diverse urban population. Leveraging the natural experiment of school closures, reopening, and mask mandates alongside changes in clinical practice that increased testing, we demonstrate the importance of viral transmission in pediatric asthma epidemiology and the increase in severe presentations of pediatric asthma in the post-COVID-19 period. Although widely observed, the post-pandemic increase in pediatric asthma severity is incompletely understood, and future work on pediatric asthma should consider this in the context of the mixtures of dynamic seasonal and static environmental risks.

## Data Availability

All data produced in the present study are available upon reasonable request to the authors, subject to institutional policies.

---

**Supplementary figure 1:**
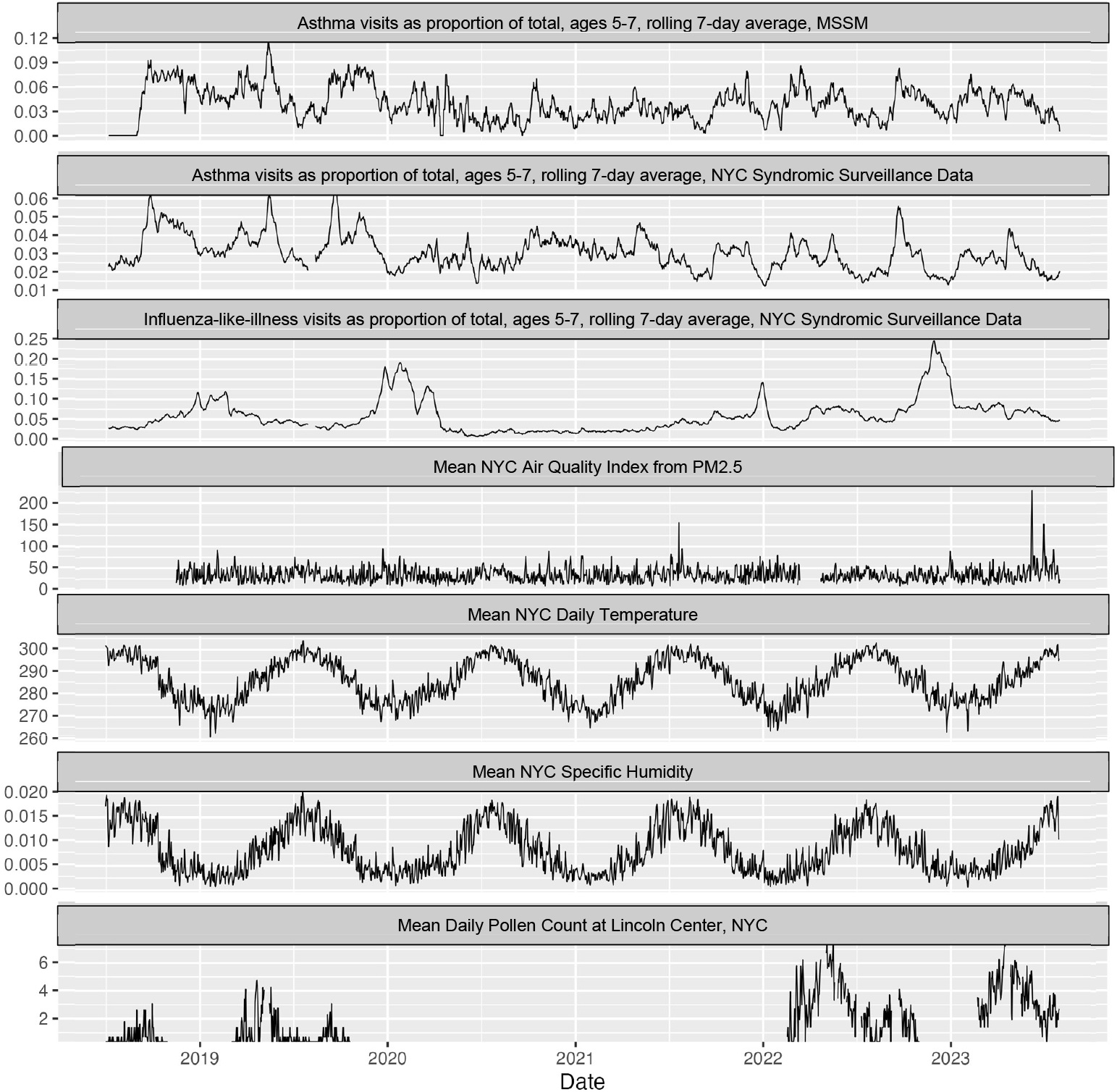
Temporal trends for asthma visits, viral (influenza-like-illness), and selected environmental exposures.

**Supplementary figure 2:**
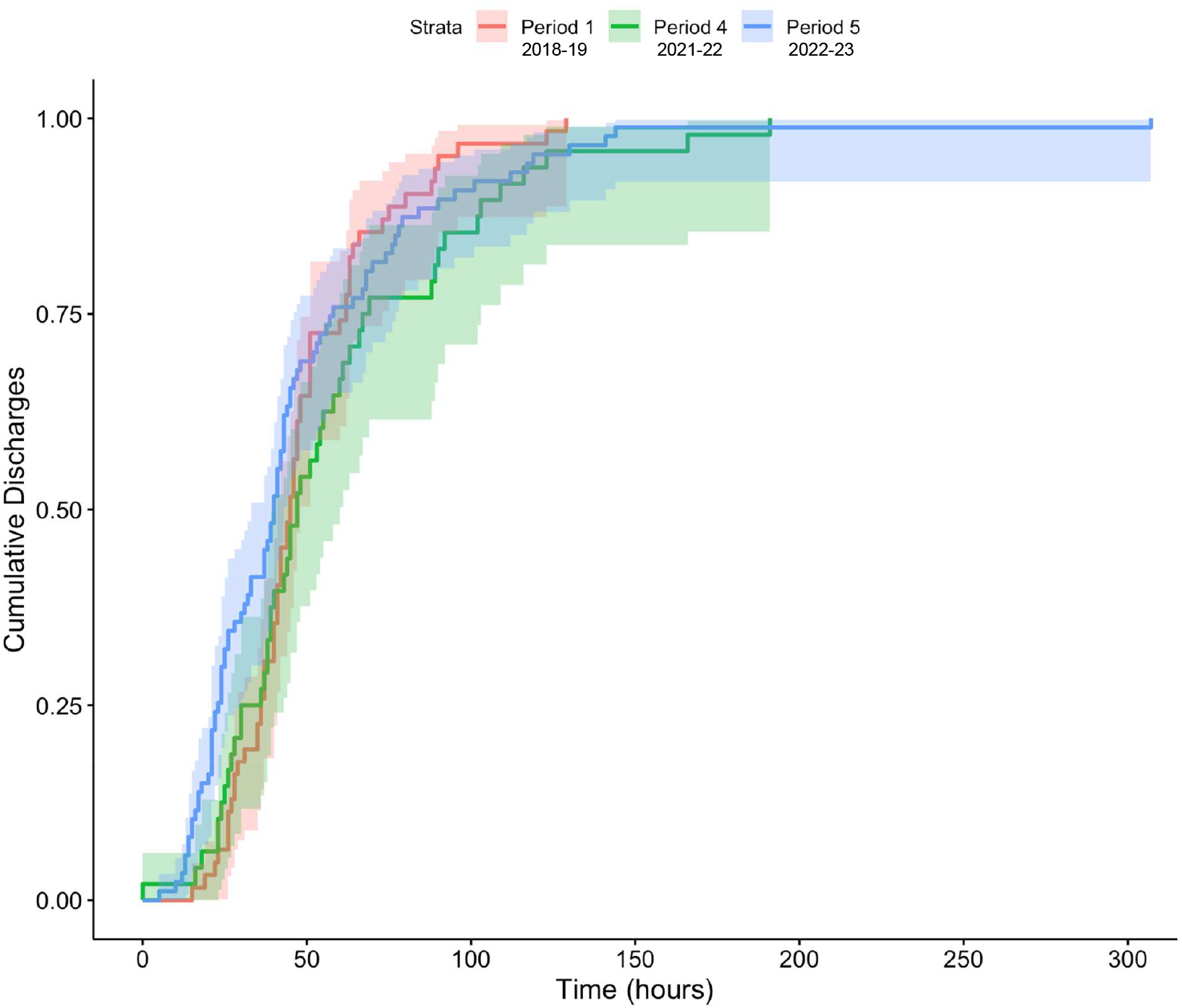
Length of stay by school year, for asthma hospital admissions, MSSM, ages 5-17 years.

